# Exploring genetic confounding of the associations between screen time and depressive symptoms in adolescence and early adulthood

**DOI:** 10.1101/2024.12.02.24318295

**Authors:** Jiayao Xu, Jessie Baldwin, Amanda M. Hughes, Annie Herbert, Hannah J. Jones, Marcus R. Munafo, Laura D. Howe

**Author notes:** Correspondence Jiayao Xu, Population Health Sciences, Bristol Medical School, University of Bristol, Bristol, BS1 5DS, United Kingdom.

## Abstract

**Background:** Digital devices have become a major aspect of children’s life. Associations between screen time and mental health have been observed, but the causality remains unclear. This study aimed to investigate the associations between screen time and later depressive symptoms, and to test the robustness of these associations when accounting for genetic confounding.

**Methods:** This study used data from the Avon Longitudinal Study of Parents and Children (ALSPAC), a prospective cohort of children born between 1991 and 1992 in the UK. Different forms of screen time and depressive symptoms at ages 16, 22 and 26 were assessed through self-completion questionnaires. Average daily screen time was calculated. Depressive symptoms were measured using the Short Mood and Feelings Questionnaire (SMFQ). Polygenic scores for depression were calculated. Linear regression models were used to examine the associations between standardised screen time at ages 16, 22 and 26 and depressive symptoms at age 26, adjusting for sociodemographic confounders and polygenic scores. Genetic sensitivity analysis (Gsens) was used to test for genetic confounding in these associations.

**Results:** A total of 3,003 participants were included in analysis. Some, but not all, forms of screen time were associated with higher SMFQ scores, e.g. time spent using a phone, tablet, or e-book at age 22 (β: 0.10 [95%CI 0.07, 0.14] for weekdays; β: 0.08 [0.04, 0.11] for weekends) and television time at age 26 (β: 0.10 [95%CI 0.06, 0.14] for weekdays; β: 0.09 [0.06, 0.13] for weekends). These associations persisted after adjusting for sociodemographic confounders and polygenic scores but were attenuated in the genetic sensitivity analysis (β=0.03 [-0.01, 0.07] for the association with time spent using a phone, tablet, or e-book at age 22 on weekends; β=0.06 [0.01, 0.10]) for television time at age 26 on weekends).

**Conclusions:** For some measures of screen time, there were no associations with depressive symptoms. Where associations were seen, they were attenuated in genetic sensitivity analysis, implying genetic confounding is present in the relationship between screen time and depressive symptoms in adolescents and young adults.

**Key Messages:**

- This study investigated the associations between screen time and later depressive symptoms during adolescence and early adulthood, using a range of approaches to examine confounding, including a genetic sensitivity model.
- Genetic confounding plays a role in the relationship between different types of screen time and depressive symptoms in adolescents and young adults, with the relationship between excessive screen time and depressive symptoms appearing weaker after accounting for genetic confounding.
- This study suggests that genetic confounding is present in the relationship between screen time and depressive symptoms in adolescents and young adults, highlighting the need to consider genetic factors when interpreting this association.

## Introduction

Digital technology is integral to children’s lives [1, 2], yet excessive use of digital devices is a global concern [3] associated with negative health outcomes, including mental health problems like depression in youth [3, 4]. Several national organisations recommend that both adolescents and young adults limit their recreational screen time [5-7], although many young people exceed the recommended amount [2, 8].

Cross-sectional and longitudinal studies have indicated that excessive screen time is associated with increased depression [8, 9]. A meta-analysis of observational studies reported dose-response associations between screen time and risk of depression in children and adolescents [10]. However, genome-wide association studies (GWAS) have indicated high genetic correlations between leisure screen time and depression [11]. Young people with a higher genetic vulnerability to depression problems are more likely to engage in screen time based on the model of compensatory internet use (that people experiencing negative moods might turn to the digital world as an escape from real life [12]), as well as be more prone to developing depressive symptoms. Therefore, the associations between screen time and depressive symptoms could be inflated by genetic confounding.

A polygenic score (PGS) represents an estimation of an individual’s genetic predisposition towards a specific trait [13, 14]. Controlling for a PGS for depression helps account for potential genetic confounding when examining the associations between screen time and depression. However, PGSs only capture a small proportion of heritability in depression, and thus do not fully account for genetic confounding [15, 16]. This limitation can be addressed by conducting a genetic sensitivity analysis, which estimates genetic confounding under scenarios in which the PGS captures additional genetic variance in depression. Specifically, this is achieved by deriving a latent PGS that captures SNP (single nucleotide polymorphisms)-based heritability in depression, and controlling for this PGS when testing the associations between screen time and depression (for more details, see refs [17, 18] and “Methods”).

Two previous studies have applied this genetic sensitivity analysis to investigate genetic confounding of the relationship between media use and mental health. First, using a cross-sectional study design with a sample of British twins assessed at age 22, Ayorech et al [19] found that the associations between media use and mental health were explained largely by genetic influences, with these findings supported by bivariate twin analyses. Second, Zhang and colleagues [20] examined the association between screen time and parent-reported psychiatric problems a year later among US children from the Adolescent Brain Cognitive Development (ABCD) study, and observed substantial genetic confounding. While these studies provide initial evidence to suggest genetic confounding, it is important to (1) test whether the findings replicate in divergent samples, with different measures of screen use, and (2) understand the extent to which genetic confounding affects longitudinal relationships between screen time and mental health, assessed several years later.

In this study, we addressed these research gaps by exploring genetic confounding in the associations between screen time and self-reported depressive symptoms using three waves of longitudinal data collected during adolescence and early adulthood. We aimed to investigate the associations between screen time and later self-reported depressive symptoms, and to test the robustness of these associations when accounting for genetic confounding, using three waves of data in adolescence and early adulthood from a UK birth cohort study.

## Methods

### Dataset

The Avon Longitudinal Study of Parents and Children (ALSPAC) is a longitudinal study that recruited about 15,000 mothers with expected dates of delivery between 1991 and 1992 in the former county of Avon, United Kingdom [21-25]. The initial number of pregnancies enrolled was 14,541 with 13,988 children alive at 1 year of age. The total sample size for analyses is 15,447 pregnancies and 14,901 children were alive at 1 year of age. For details, please refer to Supplementary Section 1.

### Inclusion and exclusion criteria

We included ALSPAC children who were alive at one year of age, responded to at least 50% of the screen time questions across the assessments at ages 16, 22 and 26, and completed at least one questionnaire on depressive symptoms at ages 16, 22 or 26 (Supplementary Table S1). For the 41 families with twins, we included the child who was born first. Participants without genotype data that passed quality control were excluded (Figure 1).

## Measurement

### Screen time

Participants’ self-reported screen time use was collected at three time points through a self-completion questionnaire during adolescence and early adulthood: at ages 16, 22, and 26. They were asked questions about screen time separately for a normal weekday and a weekend day. At age 16, they were asked about the amount of time spent watching television, using a computer, texting, and talking on a mobile phone. At ages 22 and 26, they were asked about the amount of time spent watching television, playing video games on computers/laptops or game consoles, using a computer/laptop for purposes other than gaming, and using a phone, tablet, or e-book Each response option was transformed into an ordinal variable by substituting it with the median of the corresponding category (e.g. “less than 1 hour” = 0.5) (further details of derivation and definitions are in Supplementary Table S2). Average daily screen time at each age was calculated using the formula: (total screen time on weekdays × 5 + total screen time on weekends × 2) / 7 and then standardised for comparison across the three waves.

### Depressive symptoms

Depressive symptoms were assessed using the 13-item Short Mood and Feelings Questionnaire (SMFQ) at ages 16, 22 and 26 [26]. The total score ranged from 0 to 26, where higher scores suggest more depressive symptoms. The SMFQ was reported by participants themselves at ages 16, 22 and 26 (Supplementary Table S3). The SMFQ has previously demonstrated good validity and reliability within the ALSPAC cohort, with strict measurement invariance and Cronbach’s α ranging from 0.87 to 0.92 between ages 14–26 [27, 28]. The SMFQ scores were standardised.

### Genotyping and polygenic scores

Detailed information about genotyping and quality control in the ALSPAC is provided elsewhere [29]. For the quality control process, please refer to Supplementary Section 1. PGSs for depression were generated using GWAS summary statistics [30]. PGSs were derived using PRSice software [16, 31] including SNPs regardless of their significance (p=1) [13, 16]. SNPs were clumped (with a linkage disequilibrium [LD] clumping distance of 10,000 kb and R^2^=0.01) so that the retained SNPs are largely independent and thus their effects can be summed [32, 33]. The alleles associated with the phenotype were summed and weighted by their effect sizes reported in the corresponding GWAS to compute PGSs. To control for population stratification, we residualised PGSs for the first 10 principal components estimated from the genome-wide SNP data [34]. PGSs were standardised.

### Sociodemographic factors

Sociodemographic factors were: participants’ sex at birth (females/ males), parental marital status (married/ remarried/ widowed/ single never married/ divorced/ legally separated), parental highest education levels (maternal and partner highest education qualifications (CSE / vocational/ O-level/ A-level/ degree)), parental highest occupational social classes (I/ II/ III (non-manual)/ III (manual)/ IV/ V) and not in education, employment and training (NEET) status (yes/no) (Supplementary Table S4). Parental marital status during pregnancy was combined into married/ remarried, never married, and divorced/ legally separated/ widowed. All factors, except NEET status, were reported in questionnaires completed by the child’s main caregiver (usually the mother) during pregnancy. NEET status, defined as not being engaged in employment, education or training [35], was based on self-reported information at age 16.

### Missing data

To maximize the sample size and reduce selection bias due to attrition (Supplementary Table S5), we used multiple imputation to impute missing screen time, depressive symptoms and sociodemographic data (Supplementary Table S6). Missing data were imputed using the mice package in R version 3.6.1, under a missing at random assumption. Considering sex differences in screen time and depressive symptoms and possible interactions [36], females and males were imputed separately [37]. Variables were imputed using 50 imputations with 30 iterations, the imputation model included all the variables in the analytic model (exposures, outcomes, covariates), and additional auxiliary variables that predict missingness or make the missing at random assumption more plausible (Supplementary Table S7).

### Data analyses

Descriptive analyses were conducted. Associations between each type of screen time at ages 16, 22, and 26 and depressive symptoms at age 26 were estimated using linear regression. We examined the association between screen time and depressive symptoms using an unadjusted model (Model 1), a model adjusted for sociodemographic factors (Model 2), a model adjusted for both sociodemographic factors and PGSs for depression (Model 3). The genetic sensitivity analysis was conducted using the Gsens package (Model 4) [17, 18]. After adjusting for sociodemographic factors, we assessed the association between screen time and depressive symptoms accounting for a latent PGS capturing SNP-based heritability of depression in a structural equation model (Figure 2). The genetic confounding effect is calculated as the decrease in this association after including the latent PGS. Additionally, where appropriate, we estimated the proportion of effect size attenuation attributable to genetic confounding. The SNP heritability of depression at age 26 was set at 0.05 in the Gsens model [38]. See Supplementary Section 2 for SNP-based heritability estimates and Supplementary Section 3 for the pseudo-R code.

We also conducted the analyses to examine the associations between screen time at age 16 and depressive symptoms at age 16 (SNP-based heritability=0.08), and the associations between screen time at ages 16 and 22 and depressive symptoms at age 22 (SNP-based heritability=0.05). Sensitivity analyses were conducted using heritability values ranging from 0.01 to 0.10 and using PGSs constructed with a less stringent LD clumping threshold of r^2^>0.1. Additional analysis was conducted using a binary measure of screen time. Each subtype of screen time was categorised into “> 2h per day” and “≤ 2h per day” in line with previous studies reporting a non-linear dose-response association between screen time and depression, with a cut off at two hours per day [10, 39]. Complete case analysis was also conducted as a sensitivity analysis.

## Results

### Descriptive statistics

Of 3,003 participants, 1,937 (64.5%) were females; most of them (85.9%) were from families with married parents; 1,001 (33.3%) participants’ parent/s had a degree; 77 reported NEET status at age 16 (Table 1).

The proportions of different types of screen time during weekdays and weekends are shown in Supplementary Table S2. At age 16, 34.2% of participants reported watching more than 2 hours of television, 41.8% reported using a computer for more than 2 hours, 27.1% reported texting for more than 2 hours, and 8.2% reported talking on a mobile phone for more than 2 hours during weekends. During weekdays, these proportions were lower for television and computer (24.0% and 29.7%) and similar for texting and talking on a mobile phone (21.5% and 5.6%). Proportions generally increased at age 22, and even further at 26. The median SMFQ score was 4 (interquartile range, IQR: 2 to 9) at age 16, increased to 5 (IQR: 2 to 10) at age 22 and remained at 5 (IQR: 2 to 10) at age 26. Correlations for all screen time measures are shown in Supplementary Table S8. Compared with participants excluded due to <50% data on screen time and depressive symptoms, those with ≥50% data on screen time and depressive symptoms were more likely to be female, from married or widowed families, have parents with a higher education level and be in NEET status (Supplementary Table S5).

### Associations between screen time and depressive symptoms

Table 2 shows the results of associations between each type of screen time at ages 16, 22 and 26 and SMFQ score at age 26. Unadjusted models (Model 1, Figure 3) showed that some, but not all, types of screen time were associated with the SMFQ score. Average daily screen time at ages 16 (β: 0.11 [95%CI 0.07, 0.14]), 22 (β: 0.08 [0.04, 0.12]) and 26 (β: 0.11 [0.07, 0.14]) was associated with depressive symptoms at age 26. In particular, time spent using a phone, tablet, or e-book at age 22 (β: 0.10 [0.07, 0.14] for weekdays; β: 0.08 [0.04, 0.11] for weekends) and television time at age 26 (β: 0.10 [0.06, 0.14] for weekdays; β: 0.09 [0.06, 0.13] for weekends) seem to be associated with depressive symptoms at age 26.

After adjusting for sociodemographic confounders (Model 2, Figure 3), including sex, parental marital status, parental highest education level, parental highest occupational social class and NEET status, some existing associations between screen time and depressive symptoms in the unadjusted models changed. For example, the association with texting time at age 16 (β: 0.02 [95%CI −0.02, 0.06] for weekdays; β: 0.02 [-0.01, 0.06] for weekends) attenuated. However, further adjustment for PGSs (Model 3, Figure 3) did not attenuate the observed associations.

### Associations between screen time and depressive symptoms after adjusting for latent polygenic scores capturing SNP heritability in depression

In general, most of the associations were further attenuated in the genetic sensitivity analysis models that accounted for latent PGSs for depression (Model 4, Figure 3, Table 3). The association between average daily screen time at age 16 and depressive symptoms at age 26 were strongly attenuated (β=0.04 [0, 0.08], proportion attenuation= 42%), whereas the associations at ages 22 (β=0.07 [0.03, 0.11]) and 26 (β=0.15 [0.11, 0.19]) with depressive symptoms at age 26 remained. The attenuation in the effect sizes with the genetic sensitivity analysis ranged from 13% to 55% (mean = 30%). For example, the associations between time spent using a phone, tablet, or e-book at age 22 and SMFQ scores at age 26 were as follows for weekdays and weekends: β=0.05 [0.01, 0.09], 30%; β=0.03 [-0.01, 0.07], 41%.

The effect sizes of genetic confounding in the associations between screen time and outcomes at ages 16 and 22 were similar to those for the outcome at age 26. The associations between average screen time at age 16 and depressive symptoms at ages 16 (β=0.13 [0.08, 0.17], proportion attenuation= 25%) and 22 (β=0.01 [-0.03, 0.06], proportion attenuation= 69%), and there was no evidence of genetic confounding in the associations between screen time at age 22 and depressive symptoms at age 22 (Supplementary Table S9).

Genetic confounding was similar across heritability values ranging from 0.01 to 0.10 with greater levels observed at higher heritability parameters (Supplementary Table S10). Results weren’t substantially change when using the PGS derived with an LD clumping threshold of r^2^>0.1. Additional genetic confounding was observed in the associations between average screen time at age 22 and depressive symptoms at age 26 (Supplementary Table S11). Attenuation in the associations between more than 2 hours of different types of screen time and depressive symptoms at age 26 was also observed in the genetic sensitivity models (Supplementary Table S12). Results from the complete-case analyses are presented in Supplementary Table 13.

## Discussion

Based on the large longitudinal dataset, we found some – but not all – measures of screen time were associated with higher levels of depressive symptoms. In particular, average daily screen time at age 16, time spent using a phone, tablet, or e-book at age 22 and television time at age 26 were associated with depressive symptoms at age 26. However, these associations between screen time and depressive symptoms were attenuated after accounting for genetic confounding. Due to the complex nature of genetic architecture, PGSs captured only a small proportion of total heritability [15] and were insufficient to account for total genetic confounding. Associations between screen time and depressive symptoms persisted when including PGSs as covariates but were extensively attenuated when using latent PGSs that incorporate information about SNP-based heritability. We adjusted for ancestry principal components to minimise the impact of population stratification. Nevertheless, results of the Gsens analysis might reflect several sources of genetic confounding. Besides confounding from direct genetic effects [18], this may include the impact of SNPs in LD with included SNPs and confounding due to family-level processes of assortative mating and genetic nurture [40].

Studies have suggested that genes played a role in both screen time [11] and mental health [41-43], providing potential for genetic confounding. However, most previous observational studies of mental health outcomes related to screen time [44, 45], have not considered genetic confounding. Of the two studies that have investigated genetic confounding using the Gsens analysis, one focused on screen time and psychiatric problems in children, based on one-year follow-up data [20] and the other on media use and mental health measured at the same time-point in early adulthood [19]. Both reached the similar conclusion as this paper, i.e. that genetic confounding explained most of the association between screen time and mental health [19, 20]. A PGS for depression could be associated with screen time if screen time is a consequence of, rather than a cause of, depression, or if the genetic variants affect both screen time and depression through pleiotropic pathways [11].

In this study, using a large UK longitudinal dataset with three waves of data collected during adolescence and early adulthood, we explored the associations between different types of screen time and later depressive symptoms. We investigated these associations using information on exposures and outcomes across multiple ages and applied various approaches to interrogate genetic confounding. However, several limitations should be considered. We used self-reported screen time, which can lead to recall bias and social desirability bias [46]. The collected screen time data were ordinal variables rather than continuous. However, we included all prospectively collected screen time data in ALSPAC and calculated average screen time to test the robustness of our findings across different types of screen time and at multiple time points for the same individuals. Additionally, we focused only on white participants due to the available genetic data in ALSPAC. Trans-ancestry GWASs are needed, and data on screen time, mental health, and genetic information among populations of non-European ancestry will be helpful to triangulate our findings.

## Conclusions

We found that genetic confounding is likely to partly explain the association between screen time and depressive symptoms in adolescents and young adults. There were weaker associations between different types of excessive screen time on weekdays or weekends and depressive symptoms among adolescents and young adults after accounting for genetic confounding.

## Declarations

### Ethics approval

Ethical approval for the study was obtained from the ALSPAC Ethics and Law Committee and the Local Research Ethics Committees. Informed consent for data collection and the use of data by approved researchers was obtained from participants in accordance with the recommendations of the ALSPAC Ethics and Law Committee at the time. Participants can contact the study team at any time to retrospectively withdraw consent for their data to be used. Study participation is voluntary and during all data collection sweeps, information was provided on the intended use of data. The completion of a questionnaire, either on paper or online, was considered to be written consent from participants to use their data for research purposes. Consent for the collection of biological samples was obtained in accordance with the Human Tissue Act. Full details of ethics committee approval references for ALSPAC can be found online at http://www.bristol.ac.uk/alspac/researchers/research-ethics/.

## Acknowledgements

We are extremely grateful to all the families who took part in this study, the midwives for their help in recruiting them, and the whole ALSPAC team, which includes data collection staff, data and administrations staff, technical managers and the technical staff with the Bristol Bioresource Laboratory, based within the University of Bristol.

## Author contributions

LH, JX, JB and AMH conceptualized the study design. JX, AMH, HJ and AH analysed the data. JX drafted the manuscript while LH, JB, AMH, AH, HJ, and MM revised the paper critically. All authors contributed to and have approved the final manuscript.

## Supplementary data

Supplementary data are available at *IJE* online.

## Data availability

The informed consent obtained from ALSPAC (Avon Longitudinal Study of Parents and Children) participants does not allow the data to be made available through any third party maintained public repository. Supporting data are available from ALSPAC on request under the approved proposal number, B4380. Full instructions for applying for data access can be found here: http://www.bristol.ac.uk/alspac/researchers/access/. The ALSPAC study website contains details of all available data (http://www.bristol.ac.uk/alspac/researchers/our-data/).

## Conflict of interest

None declared

## Funding

The UK Medical Research Council and Wellcome (Grant ref: MR/Z505924/1) and the University of Bristol provide core support for ALSPAC. Genomewide genotyping data was generated by Sample Logistics and Genotyping Facilities at Wellcome Sanger Institute and LabCorp (Laboratory Corporation of America) using support from 23andMe. This publication is the work of the authors and will serve as guarantors for the contents of this paper. This study was supported by the Medical Research Council (MRC) Integrative Epidemiology Unit (MC_UU_00032/7). Annie Herbert was supported by the Wellcome Trust (Grant ref: 224114/Z/21/Z).

## Use of artificial intelligence (AI) tools

No AI tools were used in the analysis of the data or in the drafting of the manuscript.

## Figure captions

**Figure 1. Flow diagram of the participants included in the analyses**

**Alt text:** Among 14,901 eligible participants, 4,251 met the criterion of having at least 50% of the information on screen time and depressive symptoms across the assessments at ages 12, 22 and 26. In total, 3,003 participants were included, after excluding 41 second-born children and 1,207 participants without genetic information.

**Figure 2. Genetic sensitivity analysis model**. Square nodes represent observed variables and oval nodes represent latent variables. Depression G refers to observed polygenic scores for depression; Depression G* refers to latent polygenic scores that capture the heritability of depression. **Alt text:** Structural equation model showing a path from a square node “Screen Time” to a square note “Depressive Symptoms”. An oval node “Depression G*” has paths to both “Screen Time” and “Depressive Symptoms” and additionally has a path to square node “Depression G”.

**Figure 3. Associations between screen time at ages 16, 22, and 26 and depressive symptoms at age 26**. Model 1: unadjusted model; Model 2: adjusted for sex, parental marital status, parental highest education level, parental highest occupational social classes and not in education, employment and training (NEET) status; Model 3: adjusted for sex, parental marital status, parental highest education level, parental highest occupational social classes, NEET status and polygenic scores for depression; Model 4: genetic sensitivity analysis using genetic sensitivity analysis (Gsens) model with latent polygenic scores for depression, adjusted for sex, parental marital status, parental highest education level, parental highest occupational social classes and NEET status.

**Alt text:** Forest plots illustrating the associations between different types of screen time and the Short Mood and Feelings Questionnaire (SMFQ) score at age 26 in Model 1, Model 2, Model 3, and Model 4. In Model 1, most types of screen time are positively associated with the SMFQ score. These associations are slightly attenuated in Model 2, remain similar in Model 3 and are further attenuated in Model 4.

## Notes

### Competing Interest Statement

The authors have declared no competing interest.

### Funding Statement

The UK Medical Research Council and Wellcome (Grant ref: 217065/Z/19/Z) and the University of Bristol provide core support for ALSPAC. This publication is the work of the authors and will serve as guarantors for the contents of this paper. This study was supported by the Medical Research Council (MRC) Integrative Epidemiology Unit (MC_UU_00032/7). Annie Herbert was supported by the Wellcome Trust (Grant ref: 224114/Z/21/Z).

### Summary of Updates

We revised the manuscript based on the peer review suggestions from the International Journal of Epidemiology.

