## Supplementary material for "Exploring genetic confounding of the associations between screen time and depressive symptoms in adolescence and early adulthood": tables

**Table 1 Sociodemographic characteristics of participants (N=3,003)**

|  | **Proportions (%)** |
| --- | --- |
| **Sex at birth** |  |
| Females | 1937(64.5) |
| Males | 1066(35.5) |
| **Parental marital status** |  |
| Married/ remarried | 2579(85.9) |
| Never married | 299(10.0) |
| Divorced/ separated/widowed | 125(4.2) |
| **Parental highest education level^a^** |  |
| CSE/ Vocational | 264(8.8) |
| O level | 673(22.4) |
| A level | 1065(35.5) |
| Degree | 1001(33.3) |
| **Parental highest occupational social classes^b^** |  |
| I | 195(6.5) |
| II | 808(26.9) |
| III (non-manual) | 779(25.9) |
| III (manual) | 635(21.1) |
| IV | 458(15.2) |
| V | 128(4.3) |
| **NEET status^c^** |  |
| Yes | 77(2.6) |
| No | 2926(97.4) |

^a^ CSEs (Certificate of Secondary Education)/ and O levels were qualifications taken at age 16 – now replaced by GCSEs (General Certificate of Secondary Education) in England, Wales and Northern Ireland. A levels are exams taken at age 18 in these countries.

^b^ Parental occupational social class was based on the higher of the mother or partner’s occupational social class using the 1991 British Office of Population and Census Statistics (OPCS) classification.

^c^ NEET status: not in education, employment and training status at age 16

**Table 2 Associations between screen time at ages 16, 22, and 26 and depressive symptom scores at age 26 (N=3,003)**

|  | **Model 1^a^** | **Model 2^b^** | **Model 3^c^** |
| --- | --- | --- | --- |
|  | **β(95%CI)** | **β(95%CI)** | **β(95%CI)** |
| **Screen time at age 16** |  |  |  |
| **Average daily screen time** | 0.11 (0.07, 0.14) | 0.06 (0.03, 0.10) | 0.06 (0.02, 0.10) |
| **Weekdays** |  |  |  |
| Television time | 0.07 (0.04, 0.11) | 0.05 (0.01, 0.08) | 0.05 (0.01, 0.08) |
| Computer time | 0.04 (0.01, 0.08) | 0.04 (0.01, 0.08) | 0.04 (0.01, 0.08) |
| Texting time | 0.06 (0.03, 0.10) | 0.02 (-0.02, 0.06) | 0.02 (-0.02, 0.05) |
| Talking time | 0.10 (0.06, 0.13) | 0.05 (0.01, 0.09) | 0.05 (0.01, 0.08) |
| **Weekends** |  |  |  |
| Television time | 0.07 (0.03, 0.10) | 0.05 (0.01, 0.08) | 0.05 (0.01, 0.08) |
| Computer time | 0.03 (-0.01, 0.06) | 0.03 (-0.01, 0.06) | 0.02 (-0.01, 0.06) |
| Texting time | 0.06 (0.03, 0.10) | 0.02 (-0.01, 0.06) | 0.02 (-0.02, 0.06) |
| Talking time | 0.07 (0.04, 0.11) | 0.03 (-0.01, 0.07) | 0.03 (-0.02, 0.06) |
| **Screen time at age 22** |  |  |  |
| **Average daily screen time** | 0.08 (0.04, 0.12) | 0.09 (0.05, 0.12) | 0.09 (0.05, 0.12) |
| **Weekdays** |  |  |  |
| Television time | 0.07 (0.04, 0.11) | 0.06 (0.03, 0.10) | 0.06 (0.03, 0.10) |
| Computer time (excluding game time) | -0.05 (-0.09, -0.02) | -0.02 (-0.06, 0.01) | -0.02 (-0.06, 0.01) |
| Game time | 0.07 (0.03, 0.10) | 0.12 (0.08, 0.15) | 0.12 (0.08, 0.15) |
| Phone time | 0.10 (0.07, 0.14) | 0.07 (0.04, 0.11) | 0.07 (0.04, 0.11) |
| **Weekends** |  |  |  |
| Television time | 0.02 (-0.01, 0.06) | 0.0002 (-0.04, 0.04) | -0.0006 (-0.04, 0.04) |
| Computer time (excluding game time) | 0.08 (0.04, 0.11) | 0.12 (0.08, 0.15) | 0.12 (0.08, 0.15) |
| Game time | 0.07 (0.03, 0.10) | 0.12 (0.08, 0.15) | 0.12 (0.08, 0.15) |
| Phone time | 0.08 (0.04, 0.11) | 0.06 (0.02, 0.09) | 0.05 (0.02, 0.09) |
| **Screen time at age 26** |  |  |  |
| **Average daily screen time** | 0.11 (0.07, 0.14) | 0.16 (0.12, 0.20) | 0.16 (0.12, 0.19) |
| **Weekdays** |  |  |  |
| Television time | 0.10 (0.06, 0.14) | 0.08 (0.05, 0.12) | 0.08 (0.04, 0.12) |
| Computer time (excluding game time) | -0.02 (-0.06, 0.01) | 0.03 (-0.004, 0.07) | 0.03 (-0.002, 0.07) |
| Game time | 0.04 (0, 0.08) | 0.13 (0.10, 0.17) | 0.13 (0.10, 0.17) |
| Phone time | 0.16 (0.12, 0.20) | 0.13 (0.10, 0.17) | 0.13 (0.09, 0.16) |
| **Weekends** |  |  |  |
| Television time | 0.09 (0.06, 0.13) | 0.07 (0.03, 0.11) | 0.08 (0.04, 0.11) |
| Computer time (excluding game time) | -0.01 (-0.04, 0.03) | 0.08 (0.04, 0.11) | 0.07 (0.03, 0.11) |
| Game time | 0.06 (0.03, 0.10) | 0.12 (0.08, 0.15) | 0.12 (0.08, 0.15) |
| Phone time | 0.12 (0.08, 0.15) | 0.10 (0.06, 0.13) | 0.10 (0.06, 0.13) |

^a^ Unadjusted model; ^b^ Adjusted for sex, parental marital status, parental highest education level, parental highest occupational social classes, not in education, employment and training (NEET) status; ^c^ Adjusted for sex, parental marital status, parental highest education level, parental highest occupational social classes, NEET status, and polygenic scores for depression

**Table 3 Associations between screen time at ages 16, 22, and 26 and depressive symptom scores at age 26 in the Gsens model (Model 4) (N=3,003)**

|  | **Association ^a^** | **Genetic confounding effect** | **Adjusted association ^b^** |
| --- | --- | --- | --- |
|  | **β(95%CI)** | **β(95%CI)** | **β(95%CI)** |
| **Screen time at age 16** |  |  |  |
| **Average screen time** | 0.06 (0.03, 0.10) | 0.03 (0, 0.05) | 0.04 (0, 0.08) |
| **Weekdays** |  |  |  |
| Television time | 0.05 (0.01, 0.08) | 0.01 (-0.01, 0.03) | 0.04 (0, 0.08) |
| Computer time | 0.04 (0.01, 0.08) | 0.02 (0, 0.05) | 0.02 (-0.02, 0.06) |
| Texting time | 0.02 (-0.02, 0.06) | 0.02 (0, 0.04) | 0 (-0.04, 0.04) |
| Talking time | 0.05 (0.01, 0.09) | 0.02 (0, 0.05) | 0.03 (-0.01, 0.07) |
| **Weekends** |  |  |  |
| Television time | 0.05 (0.01, 0.08) | 0 (-0.02, 0.02) | 0.05 (0.01, 0.09) |
| Computer time | 0.03 (-0.01, 0.06) | 0.01 (-0.01, 0.03) | 0.02 (-0.02, 0.06) |
| Texting time | 0.02 (-0.01, 0.06) | 0.02 (0, 0.04) | 0.01 (-0.03, 0.05) |
| Talking time | 0.03 (0.01, 0.06) | 0.02 (0, 0.04) | 0.01 (-0.03, 0.05) |
| **Screen time at age 22** |  |  |  |
| **Average screen time** | 0.09 (0.05, 0.12) | 0.02 (0, 0.04) | 0.07 (0.03, 0.11) |
| **Weekdays** |  |  |  |
| Television time | 0.06 (0.03, 0.10) | 0 (-0.01, 0.03) | 0.06 (0.02, 0.10) |
| Computer time (excluding game time) | -0.03 (-0.06, 0.01) | -0.01 (-0.03, 0.01) | -0.01 (-0.06, 0.02) |
| Game time | 0.12 (0.08, 0.15) | 0.02 (0, 0.04) | 0.10 (0.06, 0.14) |
| Phone time | 0.07 (0.04, 0.11) | 0.02 (0, 0.04) | 0.05 (0.01, 0.09) |
| **Weekends** |  |  |  |
| Television time | 0 (-0.04, 0.04) | 0.01 (-0.01, 0.03) | -0.01 (-0.05, 0.03) |
| Computer time (excluding game time) | 0.12 (0.08, 0.15) | 0 (-0.01, 0.03) | 0.11 (0.07, 0.15) |
| Game time | 0.12 (0.08, 0.15) | 0.01 (-0.01, 0.03) | 0.10 (0.07, 0.14) |
| Phone time | 0.06 (0.02, 0.09) | 0.02 (0, 0.04) | 0.03 (-0.01, 0.07) |
| **Screen time at age 26** |  |  |  |
| **Average screen time** | 0.16 (0.12, 0.20) | 0.01 (-0.01, 0.03) | 0.15 (0.11, 0.19) |
| **Weekdays** |  |  |  |
| Television time | 0.08 (0.05, 0.12) | 0.02 (0, 0.04) | 0.06 (0.02, 0.10) |
| Computer time (excluding game time) | 0.03 (0, 0.07) | -0.02 (-0.04, 0) | 0.05 (0.01, 0.09) |
| Game time | 0.13 (0.10, 0.17) | 0 (-0.02, 0.02) | 0.14 (0.10, 0.17) |
| Phone time | 0.13 (0.10, 0.17) | 0.02 (0, 0.04) | 0.11 (0.07, 0.15) |
| **Weekends** |  |  |  |
| Television time | 0.08 (0.04, 0.11) | 0.02 (0, 0.04) | 0.06 (0.01, 0.10) |
| Computer time (excluding game time) | 0.07 (0.03, 0.10) | 0.01 (-0.01, 0.03) | 0.06 (0.02, 0.10) |
| Game time | 0.12 (0.08, 0.15) | 0.02 (0, 0.04) | 0.10 (0.06, 0.14) |
| Phone time | 0.10 (0.06, 0.13) | 0.02 (0, 0.04) | 0.07 (0.03, 0.12) |

**^a^** Adjusted for sex, parental marital status, parental highest education level, parental highest occupational social classes, not in education, employment and training (NEET) status; ^b^ Additionally adjusted for latent polygenic scores for depression in genetic sensitivity analysis (Gsens) model.
