## Supplementary figures and images for "Exploring genetic confounding of the associations between screen time and depressive symptoms in adolescence and early adulthood"

### Figure 1

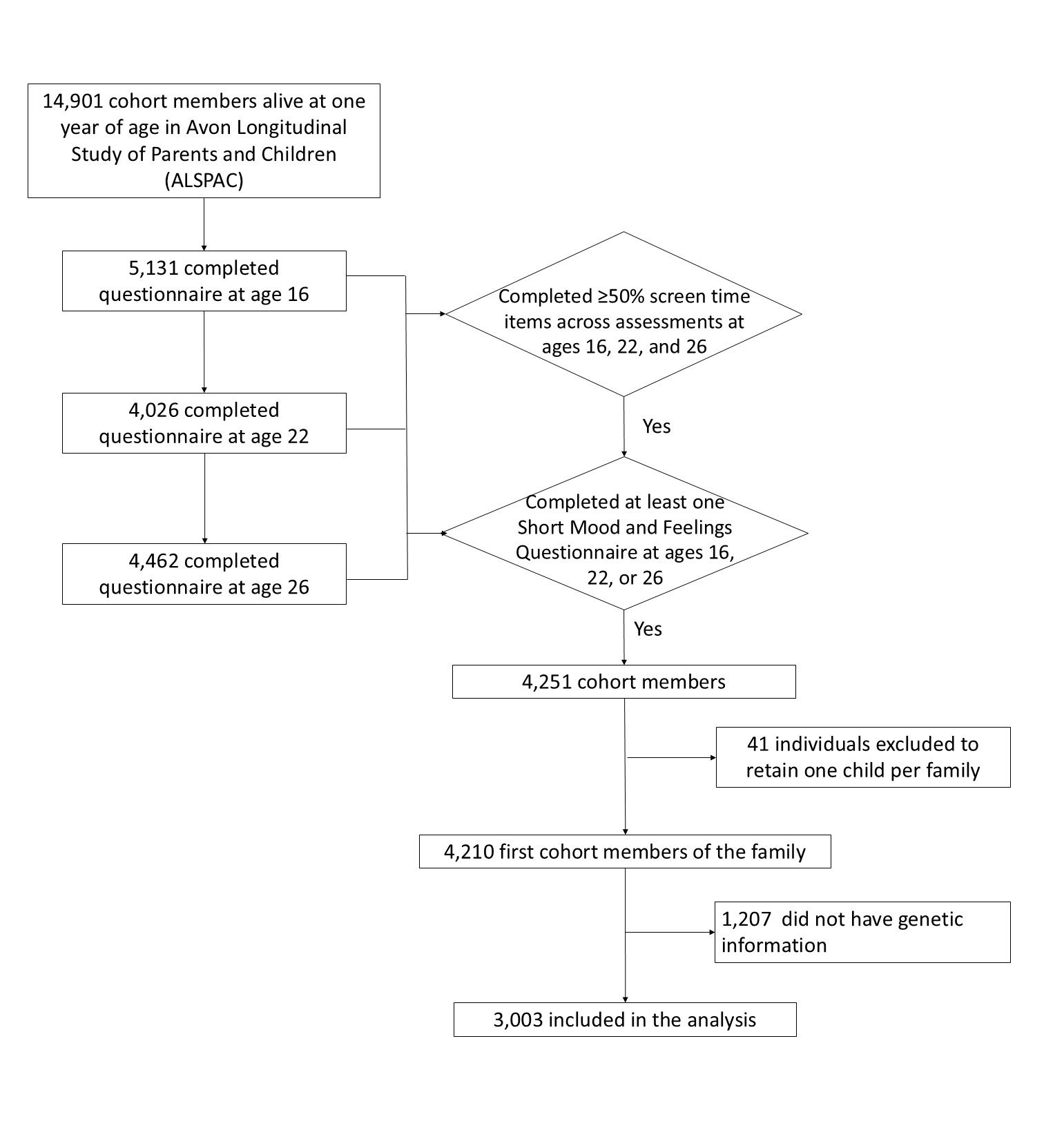

### Figure 2

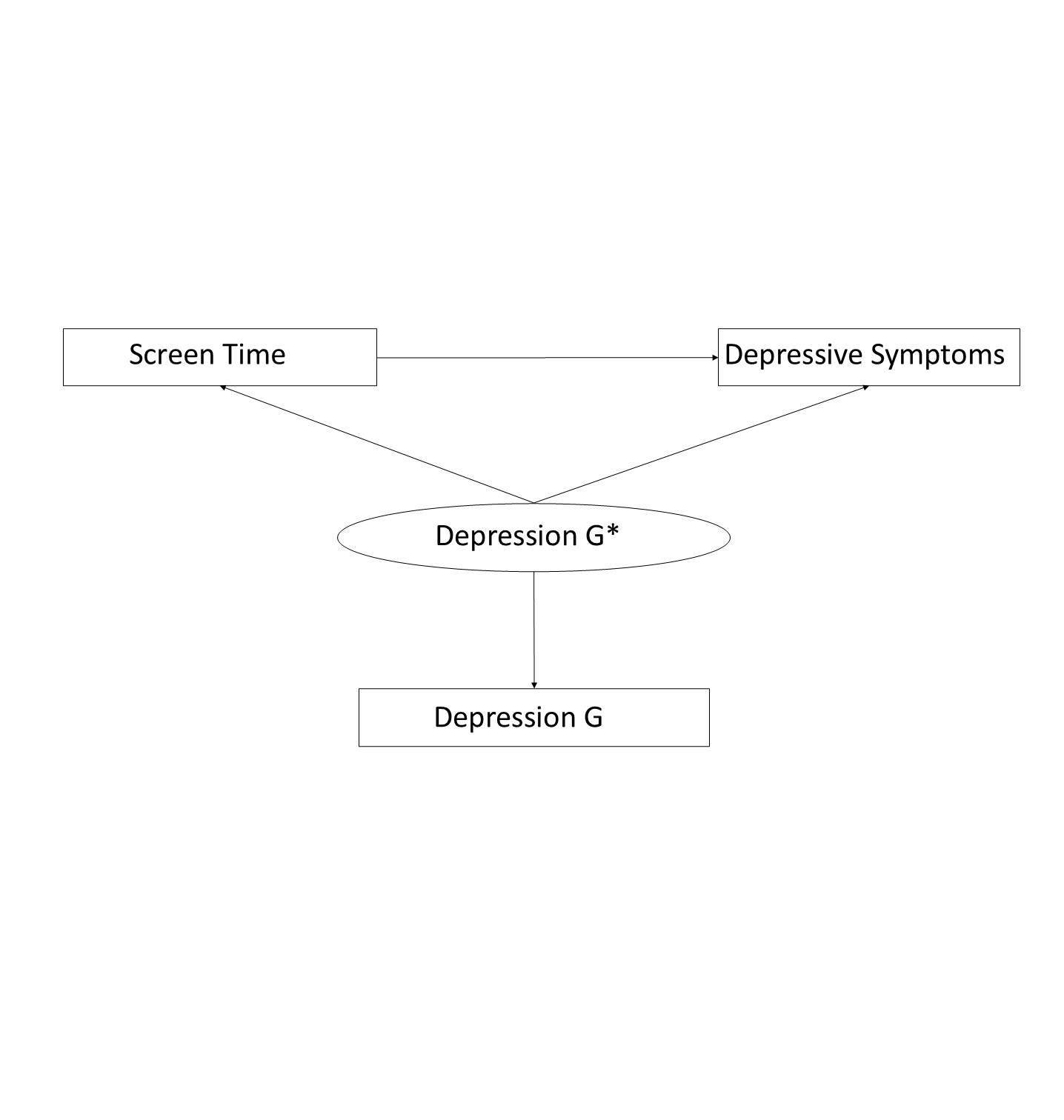

### Figure 3

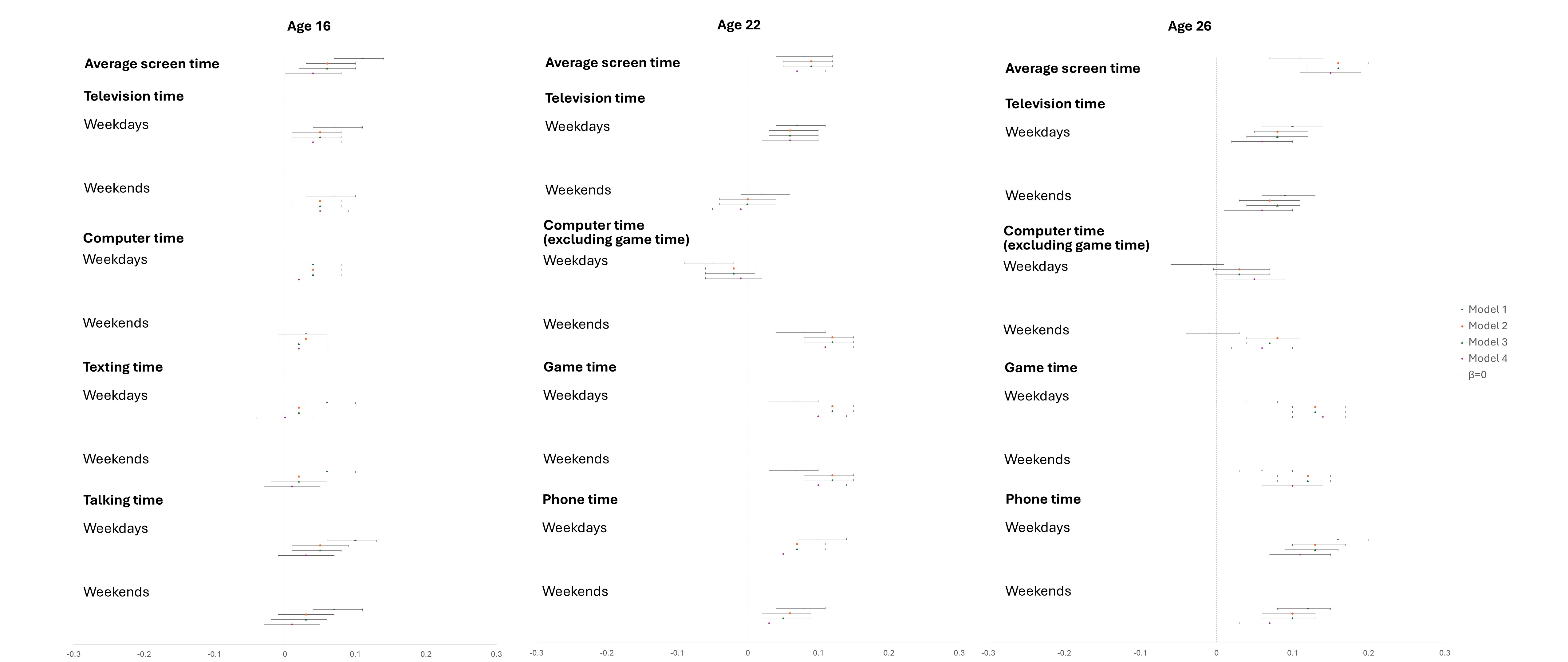
